# A Unified Framework for Statistical Inference and Study Design of Comparative *F*_1_ and *F_β_* Scores under Paired Classifier Evaluations

**DOI:** 10.64898/2026.07.15.26358166

**Authors:** Chih-Yuan Hsu, Qi Liu, Yu Shyr

**Affiliations:** Department of Biostatistics, Vanderbilt University Medical Center, Nashville, TN 37203, USA; Center for Quantitative Sciences, Vanderbilt University Medical Center, Nashville, TN 37203, USA

**Keywords:** Artificial intelligence, *F*_1_ score, *F_β_* score, classifier comparison, power analysis, sample size determination, validation study

## Abstract

**Background and objective:** The *F*_1_ score and its generalized *F_β_* score are widely used to evaluate machine learning and artificial intelligence (AI) models in healthcare, particularly for imbalanced clinical datasets. In practice, competing prediction models are commonly evaluated on the same patient cohort, resulting in correlated classifier decisions. However, existing approaches for statistical inference of *F*_1_ -related metrics typically assume independent classifier decisions or lack integrated procedures for comparative evaluation, power analysis, and sample size determination in paired validation studies.

**Methods:** We propose psF1pair, a unified framework for confidence interval estimation, hypothesis testing, and power and sample size calculation for comparative *F*_1_ and *F_β_* scores under paired evaluation designs. Dependence between classifiers is modeled using a four-component multinomial representation of the joint decision process, allowing explicit estimation and incorporation of classifier correlations commonly encountered when AI models are evaluated on the same patient cohort. Exact distributions are used for small sample settings, while asymptotic approximations are employed for computational efficiency in large studies.

**Results:** Simulation studies demonstrated that the proposed confidence intervals achieved nominal coverage probabilities across a wide range of sample sizes and correlation settings. Estimated power closely agreed with empirical power, with discrepancies generally below 3%. Compared with existing methods, psF1pair showed competitive or superior statistical power while maintaining appropriate type I error rates across a broad range of scenarios. Applications to skin cancer classification and breast cancer screening demonstrated that accounting for classifier correlation produced narrower confidence intervals and improved statistical efficiency.

**Conclusions:** psF1pair provides a practical and rigorous framework for evaluation and study planning of medical AI systems using *F*_1_ and *F_β_* metrics. The method supports comparative benchmarking, uncertainty quantification, and sample size determination for future validation studies. An open-source R package is freely available.

**Highlights:**

- Unified statistical inference and study-design framework for comparative *F*_1_ and *F_β_* scores under paired evaluation designs.
- Explicit modeling of dependence between classifiers evaluated on the same dataset using a multinomial framework.
- Open-source R package supporting comparative evaluation and validation studies of predictive and AI models.

## 1 Introduction

Machine learning and artificial intelligence (AI) systems have demonstrated substantial potential in healthcare by modeling complex clinical data and supporting diagnostic and prognostic decision-making [1]. They have become increasingly integrated into clincal practice, with applications spanning heart disease [2,3], skin and lung cancers [4,5], COVID-19 [6,7], and complication and readmission following spine surgery [8]. As AI systems become more deeply embedded in clinical workflows, rigorous evaluation of their predictive and classification performance is critical to ensure their reliability, generalizability, and clinical utility. In particular, comparative evaluation studies are increasingly conducted to determine whether newly developed AI systems outperform existing algorithms, clinical experts, or standard-of-care approaches.

The *F*_1_ score, accuray, and the area under the ROC curve (AUC) are among the most widely used metrics for evaluating diagnostic accuracy and predictive performance. The *F*_1_ score and its generaliztion, the *F_β_* score, are defined as the harmonic mean of precision (also known as positive predictive value) and sensitivity (also known as recall). Therefore, they can provide a more balanced assessment than accuracy and AUC, particularly in highly imbalanced datasets that are common in medical applications [8,9]. However, the *F*_1_ and *F_β_* scores are typically reported only as point estimates [8,10], with limited methods available for quantifying uncertainty, conducting formal comparisons between competing models, or performing power and sample size calculations for validation studies.

In recent years, there has been growing interest in developing statistical methodologies for uncertainty quantification and comparative evaluation using the *F*_1_ score. For interval estimation, Goutte and Gaussier [11] proposed a Bayesian credible interval for the *F*_1_ score, while Lam, Gopal, and Qian [12] introduced four frequentist confidence intervals (CIs) based on exact and asymptotic distributions. For hypothesis testing, Goutte and Gaussier [11] also used the same Bayesian framework to compare *F*_1_ scores under the assumption of independent classifier decisions. In contrast, Takahashi et al. [13] developed a large-sample-based method for comparing *F*_1_ scores when classifiers are evaluated on the same dataset and their decisions may be correlated. Their apparoch reduces to the independent setting when the covariance term is set to zero.

Recently, we proposed psF1 [14], a framework for interval estimation, hypothesis testing, and power and sample size calculations for single and comparative *F*_1_ and *F_β_* scores under independent classifier decisions. The framework provides practical tools for evaluating predictive models and planning validation studies, and supports exact inference for small samples and asymptotic approximations for large studies. However, the assumption of independent classifier decisions is often violated in practical AI evaluation studies.

Comparative evaluation has become a central component of medical AI research because newly developed algorithms are typically benchmarked against existing models, clinicians, or consensus interpretations using the same patient cohort. Such paired evaluation designs induce correlations between classifier decisions that should be incorporated into statistical inference and study planning. Failure to account for these correlations may lead to suboptimal confidence interval estimation, reduced statistical power, and inaccurate sample size determination. To address these challenges, we develop psF1pair, a unified framework for confidence interval construction, hypothesis testing, and power and sample size calculation for comparative *F*_1_ and *F_β_* scores in paired classifier evaluations. By explicitly modeling dependence between classifiers and providing both exact and asymptotic inference procedures, psF1pair enables rigorous validation and benchmarking of machine learning and AI systems in biomedical research. The proposed methods are implemented in an open-source R package to facilitate adoption in AI benchmarking, validation, and study-design applications.

## 2 Methods

The *F*_1_ score is defined as the harmonic mean of precision *p_p_* and sensitivity *p_s_*, given by *F*_1_ = *F*_1_(*p_s_*, *p_p_*) = 2*p_s_p_p_*/(*p_s_* + *p_p_*) . Let *N* denote the total number of class instances, with *S* positives and *N* − *S* negatives. The *F*_1_ score can be estimated as 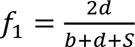[12,15], where *d* and *b* denote the numbers of true positives and false positives, respectively. Accordingly, the estimated *F*_1_ scores for classifiers 1 and 2 are *f*_1,1_ = 2*d*_1_/(*b*_1_ + *d*_1_ + *S*) and *f*_1,2_ = 2*d*_2_/(*b*_2_ + *d*_2_ + *S*), respectively, with their difference given by Δ*f*_1_ = *f*_1,1_ − *f*_1,2_.

For a common dataset of *N* instances is used, let ***Y****_i_* = (*Y*_1*i*_, *Y*_2*i*_)*^T^* denote the binary decisions of the two classifiers on the instance *i* (*i* = 1, …, *N*), where *Y*_1*i*_, *Y*_2*i*_ ∈ {0,1}. We assume independence across instances (i.e., ***Y****_i_* is independent of ***Y****_i_*′ for *i* ≠ *i*^′^), while allowing dependence between the two classifiers on the same instance (i.e., *Y*_1*i*_ may be correlated with *Y*_2*i*_). Let *E*(***Y****_i_*) = (*p*_1_, *p*_2_)*^T^*, and let *ρ* denote the correlation between *Y*_1*i*_ and *Y*_2*i*_. The joint distribution of ***Y****_i_* can be represented by a four-component indicator vector (*x*_00_, *x*_01_, *x*_10_, *x*_11_)*^T^*, generated from a one-trial multinomial distribution with cell probabilities (*π*_00_, *π*_01_, *π*_10_, *π*_11_)*^T^* . Specifically, 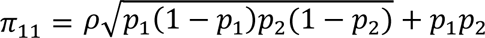, *π*_10_ = *p*_1_ − *π*_11_, *π*_01_ = *p*_2_ − *π*_11_, and *π*_00_ = 1 − *π*_10_ − *π*_01_ − *π*_11_. Under this construction, ***Y****^T^* takes values (0, 0), (0, 1), (1, 0), or (1, 1) when *x*_00_ = 1, *x*_01_ = 1, *x*_10_ = 1, or *x*_11_ = 1, respectively. An explicit form of the joint distribution of ***Y****_i_* is provided in the Supplementary Materials.

For negative instances, we assume (*p*_1_, *p*_2_)*^T^* = (*p_α_*_1_, *p_α_*_2_)*^T^* with correlation *ρ* = *ρ_N_*; for positive instances, (*p*_1_, *p*_2_)*^T^* = (*p_s_*_1_, *p_s_*_2_)*^T^* with correlation *ρ* = *ρ_P_* . Here, *p_αk_* and *p_sk_* denote the false positive rate and true positive rate of classifier *k*, for *k* = 1, 2.

Let ***b*** = (*b*_1_, *b*_2_)*^T^* denote the numbers of false positives (i.e., the sums of ***Y****_i_* over negative instances) for the two classifiers and let ***d*** = (*d*_1_, *d*_2_)*^T^* denote the numbers of true positives (i.e., the sums of ***Y****_i_* over positive instances). Then ***b*** follows a joint distribution with binomial marginals *Bin*(*N* − *S*, *p_α_*_1_) and *Bin*(*N* − *S*, *p_α_*_2_), with correlation *ρ_N_*. Similarly, ***d*** follows a joint distribution with binomial marginals *Bin*(*S*, *p_s_*_1_) and *Bin*(*S*, *p_s_*_2_), with correlation *ρ_P_*. We denote the two joint distributions by *B*_2_(***b***; *N* − *S*, *p_α_*_1_, *p_α_*_2_, *ρ_N_*) and *B*_2_(***d***; *S*, *p_s_*_1_, *p_s_*_2_, *ρ_P_*), respectively with explicit forms given in the Supplementary Materials. In the special case where *ρ_N_* = 0 and *ρ_P_* = 0, these reduce to independent binomial products: *B*_2_(***b***; *N* − *S*, *p_α_*_1_, *p_α_*_2_, 0) = *Bin*(*b*_1_; *N* − *S*, *p_α_*_1_)*Bin*(*b*_2_; *N* − *S*, *p_α_*_2_) and *B*_2_(***d***; *S*, *p_α_*_1_, *p_α_*_2_, 0) = *Bin*(*d*_1_; *S*, *p_s_*_1_)*Bin*(*d*_2_; *S*, *p_s_*_2_).

In practice, study designs are often specified in terms of target precision levels *p_pk_* rather than false positivity rates *p_αk_*. Following psF1 [14], we map precision levels to the corresponding false positive rates. Specifically, conditional on *S* and *N* − *S*, and given *p_p_*_1_, *p_p_*_2_, *p_s_*_1_, *p_s_*_2_, the *p_αk_* is the solution to

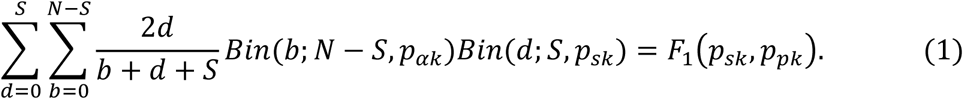

Therefore, the support of Δ*f*_1_ can be obtained by enumerating all possible combinations of ***b*** and ***d***, with probabilities determined by their joint distributions. The probability mass function of Δ*f*_1_ = Δ*f* is given by

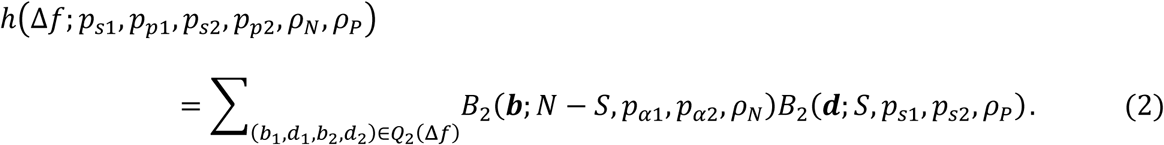

Here, *Q*_2_(Δ*f*) = {(***b***, ***d***): *f*_1,1_ − *f*_1,2_ = Δ*f*, *d_k_* ∈ ℤ*_S_*, *b_k_* ∈ ℤ*_N_*_−*S*_, *k* = 1, 2}, Δ*f* ∈ Δℱ*_N_*_,*S*_ = {*f*_1,1_ − *^f^*1,2 ^: *f*^1,1 ^, *f*^1,2 ^∈ ℱ^*N*,*S*}, 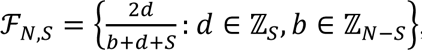 and ℤ*_n_* = {0,1, …, *n*}.

### 2.1 Confidence interval

To construct a confidence interval for the difference in *F*_1_ scores between two classifiers, the parameters *p_s_*_1_, *p_p_*_1_, *p_s_*_2_, *p_p_*_2_, *ρ_N_*, and *ρ_P_* need to be estimated. Then, substituting these parameter estimates into the distribution of Δ*f*_1_ yields a 100(1 − *α*^∗^)% confidence interval for the difference in *F*_1_ scores, where 1 − *α*^∗^ is a pre-specified confidence level. Specifically, given observed values *b_k_* = *b_obs_*_,*k*_ and *d_k_* = *d_obs_*_,*k*_ for *k* = 1, 2, the interval estimate for Δ*f*_1_ is denoted by [*L*, *U*], where

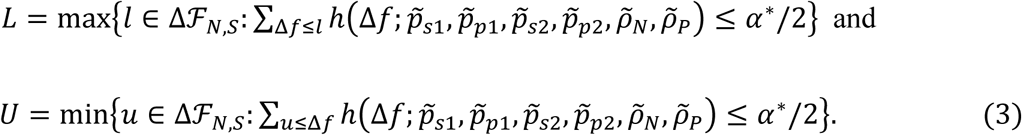

Generally, the plug-in estimates are *p̂_sk_* = *d_obs_*_,*k*_/*S* and *p̂_pk_* = *d_obs_*_,*k*_/(*b_obs_*_,*k*_ + *d_obs_*_,*k*_), along with 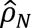 and 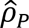, obtained as the Pearson correlation coefficient estimates of ***Y****_i_* within the negative and positive instances, respectively. However, when *N* and *S* are small, the interval estimate may exhibit insufficient coverage probability, especially when *b_obs_*_,*k*_ = 0. To mitigate this issue in small-sample settings, we replace *b_obs_*_,*k*_ with 0.5 if *b_obs_*_,*k*_ = 0, and consider *p̃_sk_* = (*d_obs_*_,*k*_ + 0.5)/(*S* + 1) and *p̃_pk_* = (*d_obs_*_,*k*_ + 0.5)/(*b_obs_*_,*k*_ + *d_obs_*_,*k*_ + 1) for *k* = 1, 2. These adjusted estimates equal the posterior means of *p_sk_* and *p_pk_* under Jeffrey’s non-informative prior [11]. The 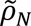 and 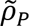 are taken as Bayesian Pearson correlation estimates obtained using a Dirichlet prior with Jeffrey’s non-informative parameters (see Supplementary Materials for details).

### 2.2 Hypothesis testing

To test *H*_0_: *F*_1,1_ = *F*_1,2_ vs. *H*_1_: *F*_1,1_ ≠ *F*_1,2_, we construct the null distribution of Δ*f*_1_ by assuming *F*_1,1_ = *F*_1,2_ = *F̂*_1,0_, where *F̂*_1,0_ = *F*_1_(*p̄_s_*, *p̄_p_*). Here, *p̄_s_* = (*p̂_s_*_1_ + *p̂_s_*_2_)/2 and *p̄_p_* = (*p̂_p_*_1_ + *p̂_p_*_2_)/2 denote the common sensitivity and precision, respectively, for the two classifiers. The estimates *p̂_sk_* = *d_obs_*_,*k*_/*S* and *p̂_pk_* = *d_obs_*_,*k*_/(*b_obs_*_,*k*_ + *d_obs_*_,*k*_) are used for *k* = 1, 2, and the Bayesian Pearson correlation estimates 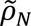 and 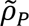 are adopted. Under this specification, the null distribution of Δ*f*_1_ = Δ*f* is given by ℎ(Δ*f*; *p̄_s_*, *p̄_p_*, *p̄_s_*, *p̄_p_*, 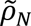, 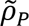) . Given the observed *F*_1_ score difference Δ*f_obs_* = *f_obs_*_,1_ − *f_obs_*_,2_, the two-sided p-value is given by

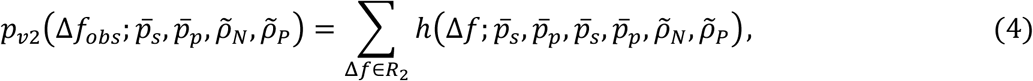

where *f_obs_*_,*k*_ = 2*d_obs_*_,*k*_/(*b_obs_*_,*k*_ + *d_obs_*_,*k*_ + *S*), *R*_2_ = {Δ*f*: *C*_2_(Δ*f*) ≤ *C*_2_(Δ*f_obs_*) }, and *C*_2_(*u*) = min{∑_Δ*f*≤*u*_ ℎ(Δ*f*; *p̄_s_*, *p̄_p_*, *p̄_s_*, *p̄_p_*, 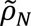, 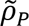), ∑_Δ*f*≥*u*_ ℎ(Δ*f*; *p̄_s_*, *p̄_p_*, *p̄_s_*, *p̄_p_*, 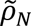, 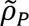)}. The set *R*_2_ consists of values of Δ*f* that are more extreme than Δ*f_obs_*. If *p_v_*_2_(Δ*f_obs_*; *p̄_s_*, *p̄_p_*, 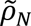, 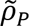) < *α*^∗^, we reject *H*_0_: *F*_1,1_ = *F*_1,2_, where *α*^∗^ is a pre-specified significance level.

### 2.3 Power and sample size calculation

Given *N*, *S*, and a pre-specified significance level of *α*^∗^, the power to detect the difference between *F*_1_(*p_s_*_1_, *p_p_*_1_) and *F*_1_(*p_s_*_2_, *p_p_*_2_), with correlations *ρ_N_* and *ρ_P_*, is given by

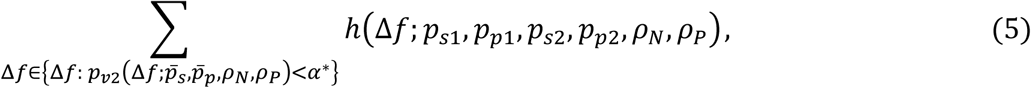

where *p̄_s_* = (*p_s_*_1_ + *p_s_*_2_)/2 and *p̄_p_* = (*p_p_*_1_ + *p_p_*_2_)/2. Based on Formula (5), the required *N* and *S* to achieve a desired statistical power at the significance level of *α*^∗^ can be determined.

### 2.4 Normal approximation

When *N* and *S* are large, the distribution of Δ*f*_1_ can be well approximated by a normal distribution. This normal approximation greatly reduces computational burden of evaluating p-values and power under exact distributions, thereby enhancing the practicality of psF1pair.

As described in Section 2, we assume independence across instances, implying that ***b*** and ***d*** are independent, and that they follow *B*_2_(***b***; *N* − *S*, *p_α_*_1_, *p_α_*_2_, *ρ_N_*) and *B*_2_(***d***; *S*, *p_s_*_1_, *p_s_*_2_, *ρ_P_*), respectively. When *N* − *S* and *S* are large, the central limit theorem implies

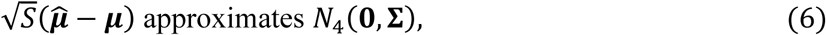

where 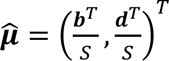, 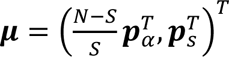 with ***p****_α_* = (*p_α_*_1_, *p_α_*_2_) and ***p****_s_* = (*p_s_*_1_, *p_s_*_2_). The covariance matrix is block-diagonal, **Σ** = bdiag(**Σ*_b_***, **Σ*_d_***), where

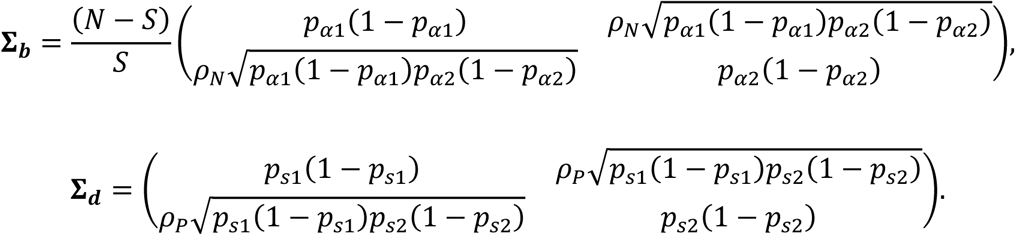

Define 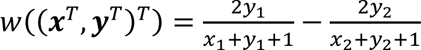 and let ∇*w*((***x****^T^*, ***y****^T^*)*^T^*) = (*∂w*/*∂**x**^T^*, *∂w*/*∂**y**^T^*)*^T^*. Then, using the Delta method, 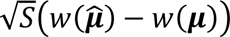) approximates *N*_1_ (0, (∇*w*(***μ***)) **Σ** ∇*w*(***μ***)). Therefore, 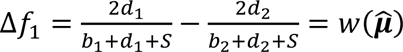 is approximately normal distributed with *E*(Δ*f*) ≈ *w*(***μ***) and *Var*(Δ*f*_1_) ≈ (∇*w*(***μ***))*^T^* **Σ** ∇*w*(***μ***)/*S*

### 2.5 Extension to comparative *F_β_* score

The proposed methods can be straightforwardly extended to comparative *F_β_* scores. The *F_β_* score is defined as:

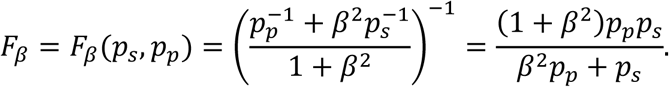

When *β* = 1, *F_β_* reduces to the standard *F*_1_ score. For *β* < 1, the metric places greater emphasis on precision, while for *β* > 1, it assigns more weight to sensitivity. In this extension, we replace Δ*f*_1_, *F*_1,1_, *F*_1,2_, and *F̂*_1,0_ with Δ*f_β_*, *F_β_*_,1_, *F_β_*_,2_, and *F̂_β_*_,0_, respectively, and redefine *p_αk_* as the solution to

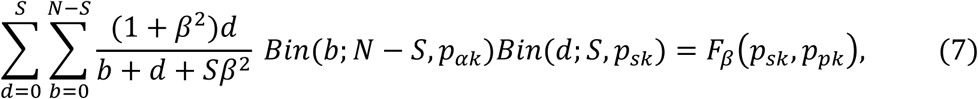

for *k* = 1, 2. Under this formulation, statistical inference and power analysis can be developed for comparative *F_β_* scores. The corresponding estimator of the comparative *F_β_* is then given by Δ*f_β_* = *f_β_*_,1_ − *f_β_*_,2_, where *f_β_*_,1_ = (1 + *β*^2^)*d*_1_/(*b*_1_ + *d*_1_ + *Sβ*^2^) and *f_β_*_,2_ = (1 + *β*^2^)*d*_2_/(*b*_2_ + *d*_2_ + *Sβ*^2^).

## 3 Validation of statistical performance

This section is organized into three subsections. First, we examined interval length and coverage probability for the *F_β_* score difference. Second, we compared psF1pair with existing hypothesis testing methods for the *F*_1_ score, as no formal testing procedures for general *F_β_* score differences are available. Third, we assessed the statistical power of psF1pair through simulation studies.

### 3.1 Confidence interval performance

We considered two scenarios to examine interval length and coverage probability of 95% CI for the *F_β_* score, with *β* = 0.5, 1, and 2, under three correlation levels *ρ_N_* = *ρ_P_* = 0, 0.2, and 0.4. In the first scenario, classifier 2 had higher precision and sensitivity than classifier 1, with (*p_p_*_2_, *p_s_*_2_) = (0.7, 0.9) compared to (*p_p_*_1_, *p_s_*_1_) = (0.6, 0.8). In the second scenario, both classifiers had identical performance, with *p_p_*_1_ = *p_p_*_2_ = 0.6 and *p_s_*_1_ = *p_s_*_2_ = 0.8. Each scenario was evaluated under four configurations: (*N*, *S*) = (250, 100), (100, 40), (50, 20), and (1000, 20), based on 1000 simulation runs.

Across all settings, the proposed 95% CIs achieved coverage probabilities close to the nominal 95% level (Figure 1). Coverage bias was more pronounced when *S* was small but improved as *S* increased. The interval length decreased with larger *S* and higher correlation levels (Figure 2).

**Figure 1.**
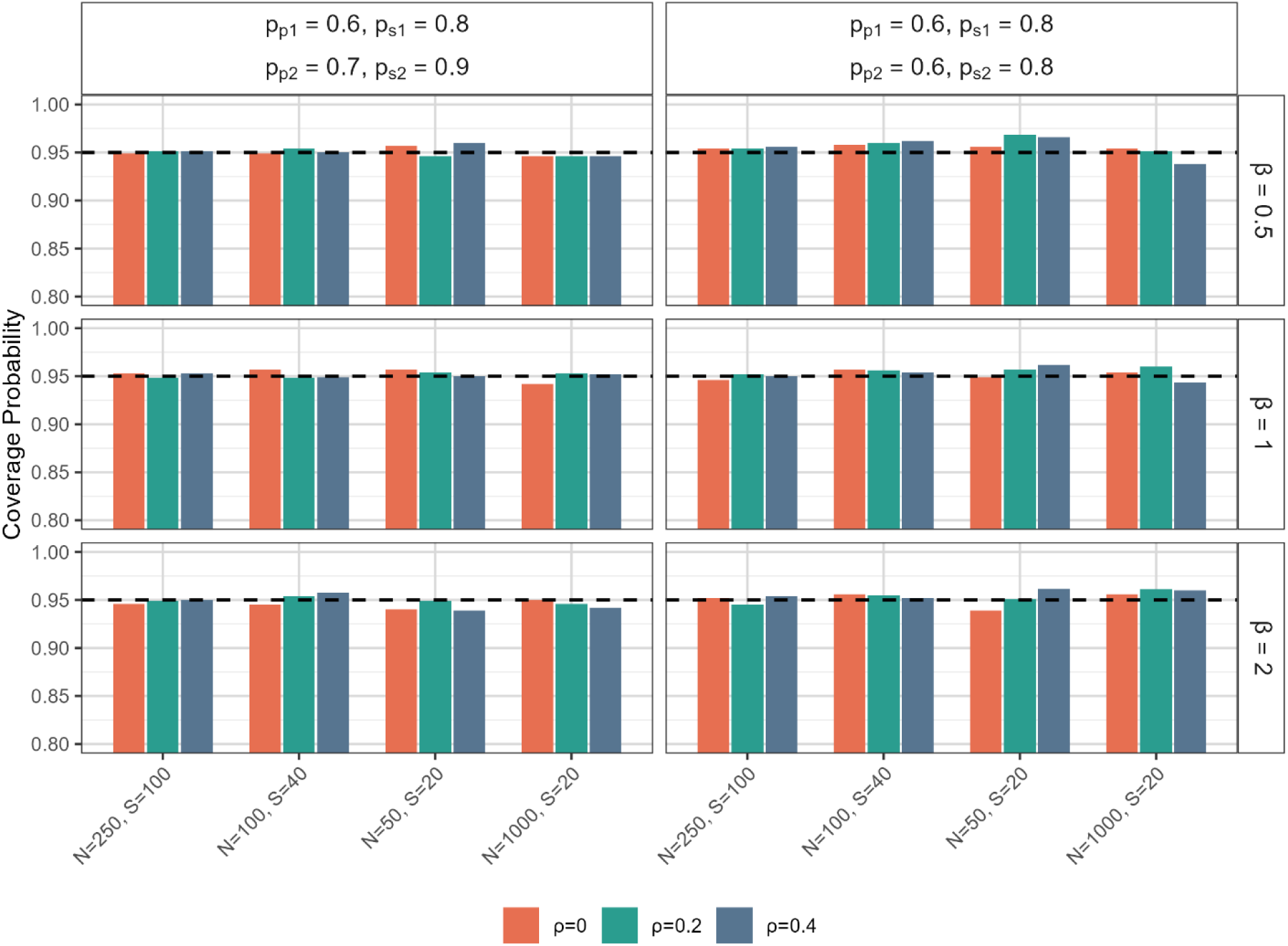
Coverage probability of 95% confidence intervals for the difference in *F_β_* scores based on 1000 simulation runs. In the first scenario (left), classifier 2 has higher prediction and sensitivity than classifier 1; in the second scenario (right), classifiers 2 and 1 have identical prediction and sensitivity. The dashed lines represent the nominal 95% level.

**Figure 2.**
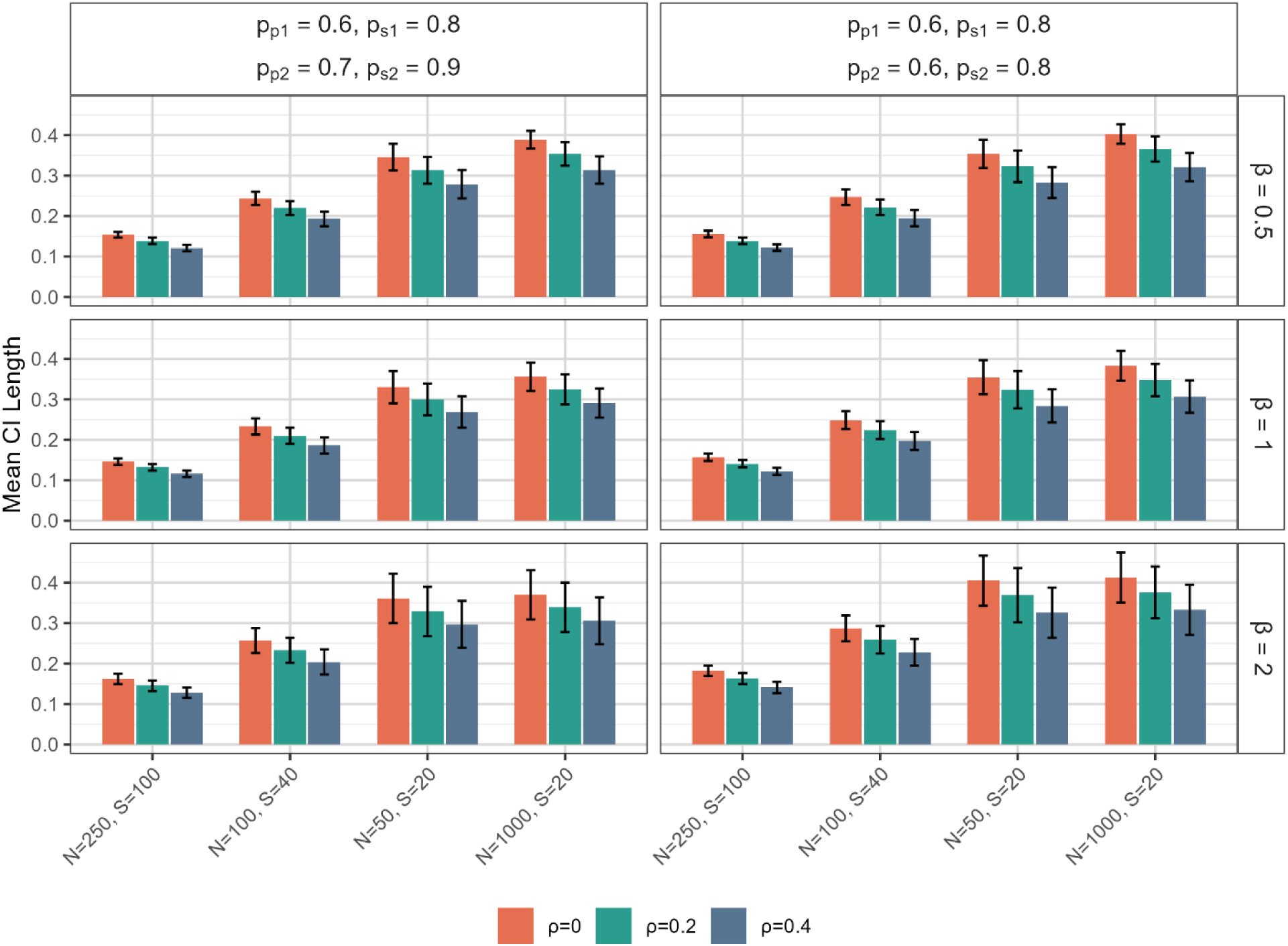
Mean length of 95% confidence intervals for the difference in *F_β_* scores based on 1000 simulation runs. In the first scenario (left), classifier 2 has higher precision and sensitivity than classifier 1; in the second scenario (right), classifiers 2 and 1 have identical precision and sensitivity. The error bars represent ± one standard deviation.

Results were similar across all three values of *β*. Detailed numerical values were provided in the Supplementary Materials (Tables S1-S3).

### 3.2 Comparative evaluation against existing methods

Five scenarios were considered: the first three for evaluating statistical power and the last two for examining type I error rates. The parameter settings were defined as follows. Compared with classifier 1, classifier 2 had: (i) higher precision and sensitivity (*p_p_*_2_, *p_s_*_2_) = (0.7, 0.9) vs. (*p_p_*_1_, *p_s_*_1_) = (0.6, 0.8), (ii) higher precision but the same sensitivity (*p_p_*_2_, *p_s_*_2_) = (0.8, 0.8) vs. (*p_p_*_1_, *p_s_*_1_) = (0.6, 0.8), and (iii) the same precision but higher sensitivity (*p_p_*_2_, *p_s_*_2_) = (0.8, 0.8) vs. (*p_p_*_1_, *p_s_*_1_) = (0.8, 0.6). Scenarios (iv) and (v) assumed identical classifier performance: (iv) lower precision and higher sensitivity (*p_p_*_1_ = *p_p_*_2_= 0.6, *p_s_*_1_ = *p_s_*_2_ = 0.8) and (v) higher precision and lower sensitivity (*p_p_*_1_ = *p_p_*_2_ = 0.8, *p_s_*_1_ = *p_s_*_2_ = 0.6). The correlations between the two classifiers among negative and positive instances were set to *ρ_N_* = *ρ_P_* = 0, 0.2, and 0.4. Each scenario was evaluated under four configurations: (*N*, *S*) = (250, 100), (100, 40), (50, 20), and (1000, 20).

psF1pair was compared with a permutation-based method (where outcomes within discordant pairs were randomly exchanged to generate an empirical null distribution of the *F*_1_ score difference) and the Wald- and score-type Takahashi methods [13], based on 1000 simulation runs. For power comparison (Figure 3, Tables S4-S6), when *ρ_N_* = *ρ_P_* = 0, psF1pair and the Wald-type Takahashi method performed best in scenario (i) and (ii), while psF1pair outperformed all other methods in scenario (iii). When *ρ_N_* = *ρ_P_* = 0.2 and 0.4, the Wald-type Takahashi method had the highest power in scenarios (i) and (ii), with the other methods performing similarly, while psF1pair remained superior in scenario (iii). For type I error examination (Figure 4, Tables S7-S9), all methods controlled well when *ρ_N_* = *ρ_P_* = 0. However, when *ρ_N_* = *ρ_P_* = 0.2 and 0.4, the Wald-and score-type Takahashi methods exhibited inflated type I error rates for (*N*, *S*) = (50, 20) and/or (1000, 20).

**Figure 3.**
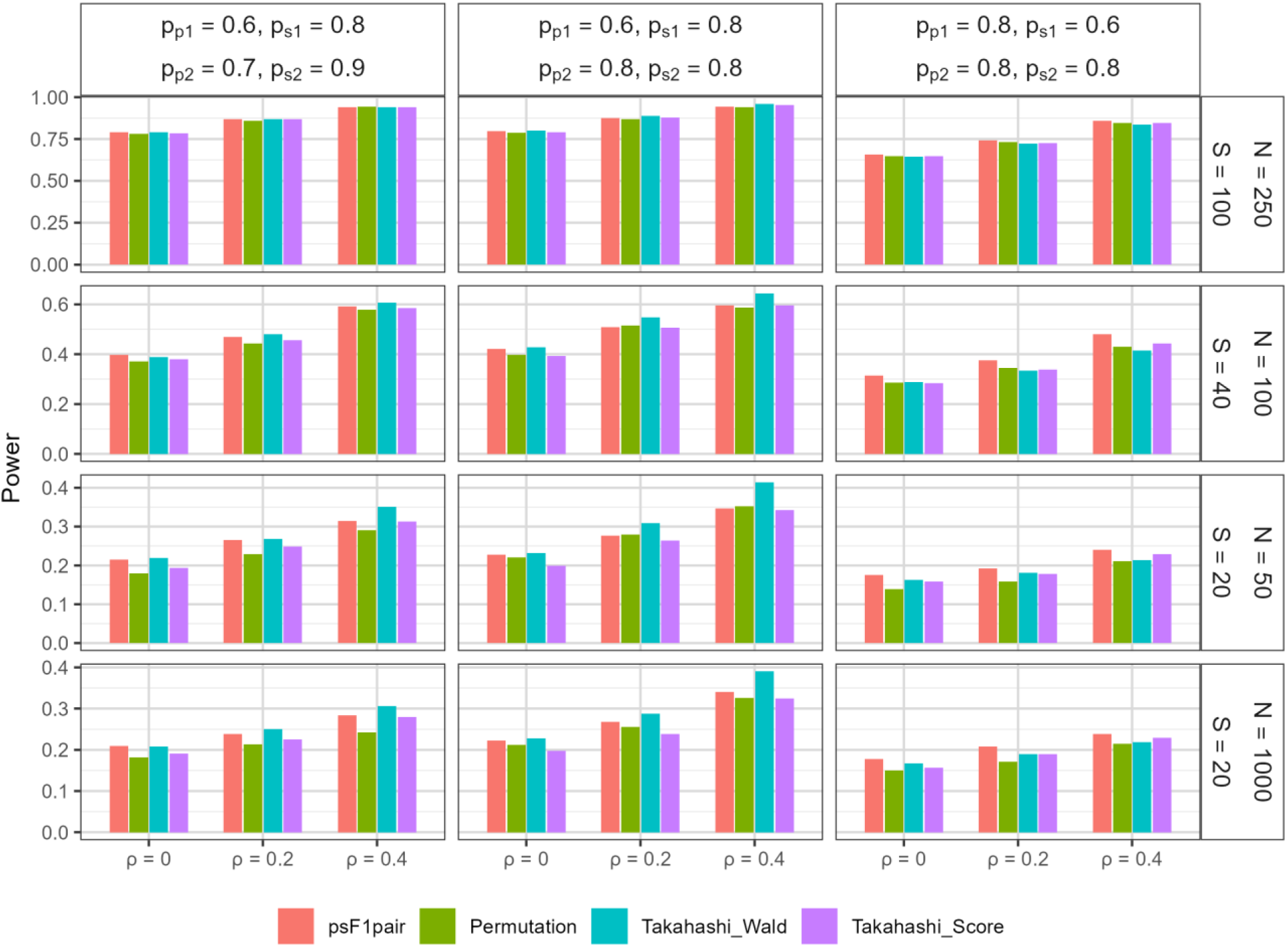
Power comparison across methods for the difference in *F*_1_ scores at the significance level of 0.05, based on 1000 simulation runs. In the first scenario (left), classifier 2 has higher precision and sensitivity than classifier 1; in the second scenario (middle), classifier 2 has higher precision but equal sensitivity compared to classifier 1; in the third scenario (right), classifier 2 has higher sensitivity but equal precision compared to classifier 1.

**Figure 4.**
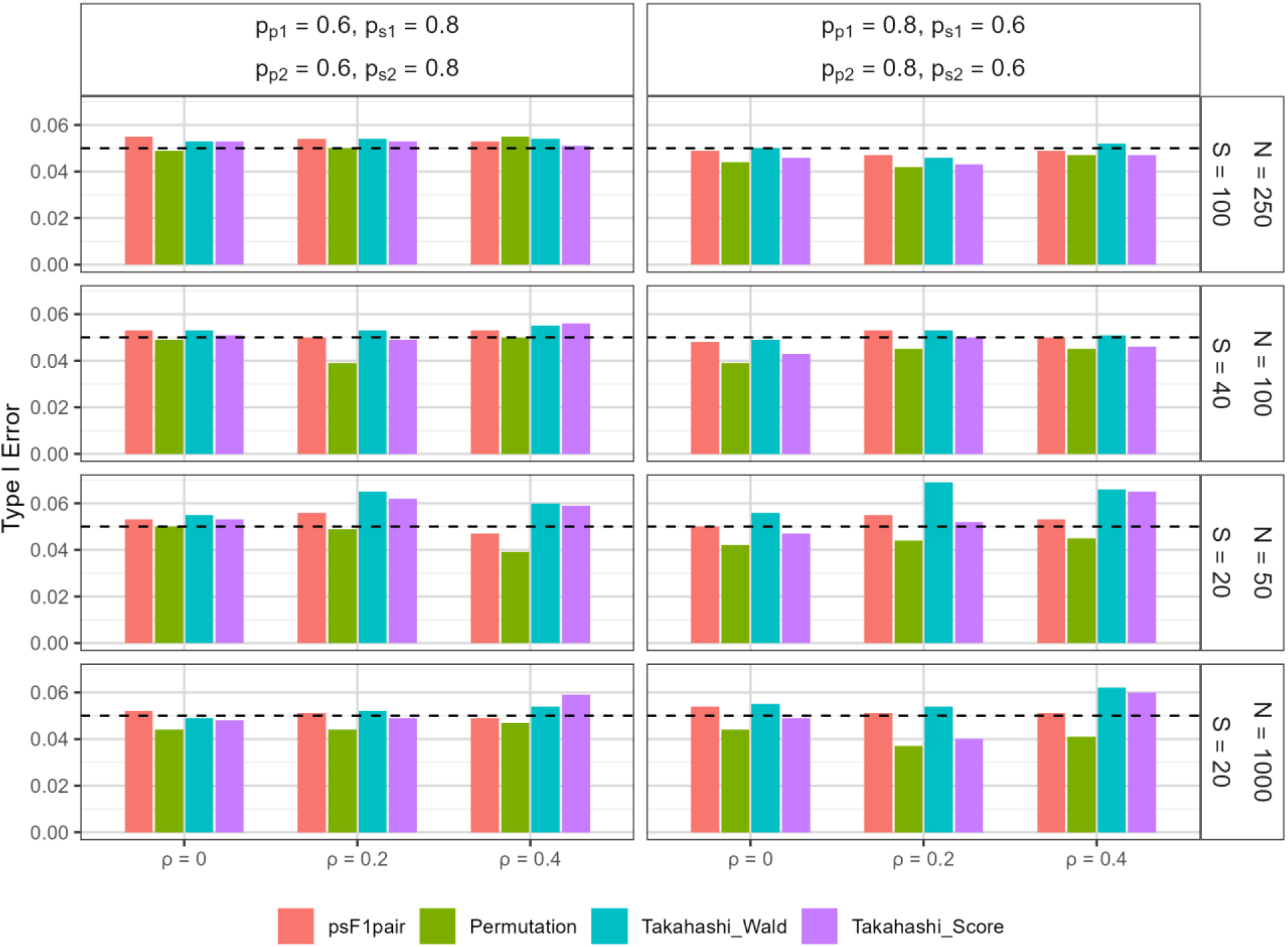
Comparison of type I error rate across methods for the difference in *F*_1_ scores at the significance level of 0.05, based on 1000 simulation runs. In both scenarios, classifiers 2 and 1 have identical precision and sensitivity. In the first scenario (left), sensitivity is higher and precision is lower, while in the second scenario (right), precision is higher and sensitivity is lower. The dashed lines represent the nominal 5% level.

### 3.3 Validation of power calculations

We assessed the accuracy of the statistical power estimated by psF1pair by comparing them with empirical power obtained from 1000 simulation runs. This validation used scenarios (i)–(iii) described in Section 3.2 for the *F_β_* score, with *β* = 0.5, 1, and 2, under three correlation levels *ρ_N_* = *ρ_P_* = 0, 0.2, and 0.4. Across all settings, the estimated powers closely matched the empirical powers, with differences of no more than 3% (Figure 5, Tables S10-S12). These results demonstrate the accuracy of psF1pair under a wide range of settings.

**Figure 5.**
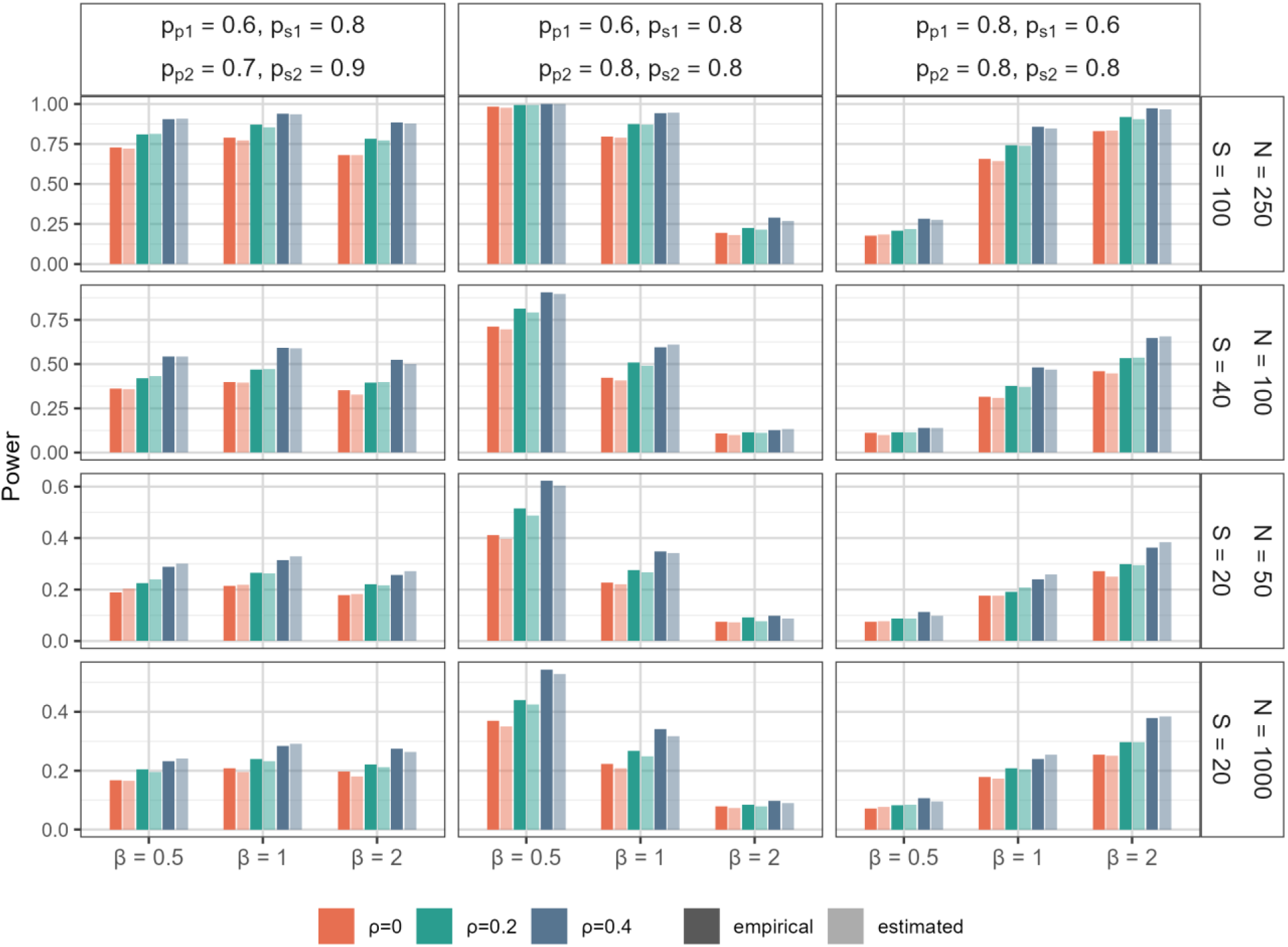
Comparison of empirical and estimated powers for the difference in *F_β_* scores at the significance level of 0.05. Empirical powers (dark bars) are based on 1000 simulation runs, while estimated powers (light bars) are calculated using psF1pair. In the first scenario (left), classifier 2 has higher precision and sensitivity than classifier 1; in the second scenario (middle), classifier 2 has higher precision but equal sensitivity compared to classifier 1; in the third scenario (right), classifier 2 has higher sensitivity but equal precision compared to classifier 1.

## 4 Real applications

We applied psF1pair to two real-world applications: an image-based skin cancer classification study [16] and a breast cancer screening study using mammograms [17].

### 4.1 Skin cancer classification

The objective of this study was to compare the diagnostic performance of a faster, region-based convolutional neural network algorithm (FRCNN) with that of board-certified dermatologists (BCD). We applied psF1pair to compare the two classification methods using the *F*_1_ score. Six skin conditions were grouped into two categories, following the classification in Takahashi et al. [13]: malignant tumors (malignant melanoma and basal cell carcinoma) and benign tumors (nevus, seborrheic keratosis, senile lentigo, and hematoma/hemangioma). The data was obtained from Table B1 in Takahashi et al. [13], which reports predictions for 2000 skin cancer images, including 540 malignant and 1460 benign cases. The classification results for malignant and benign categories are summarized in Table 1.

**Table 1.** Predicted results for malignant (M) and benign (B) tumors in the study of skin cancer classification.

|  |  | Benign (B) |  | Malignant (M) |  |  |
| --- | --- | --- | --- | --- | --- | --- |
|  |  | BCD |  | BCD |  |  |
|  |  | B | M | B | M |  |
| FRCNN | B | 1,226 | 153 | 35 | 55 |  |
|  | M | 39 | 42 | 39 | 411 |  |
| Stratified by BCD and FRCNN |  |  |  |  |  |  |
|  |  | BCD |  | FRCNN |  |  |
|  |  | B | M | B | M | Total |
| Actual | B | 1,265 | 195 | 1,379 | 81 | 1,460 |
|  | M | 74 | 466 | 90 | 450 | 740 |

For FRCNN and BCD, the estimated precisions were 0.847 and 0.705 and the estimated sensitivity were 0.833 and 0.863, respectively. The resulting *F*_1_ scores were 0.840 and 0.776, yielding a difference of 0.064. Using psF1pair, this difference was statistically significant, with a p-value = 3.76 × 10^−6^. In comparison, the Wald- and score-type Takahashi methods yielded p-values of 1.07 × 10^−5^ and 1.54 × 10^−5^, respectively.

The estimated correlation between FRCNN and BCD was 0.327 for malignant tumors and 0.274 for benign tumors. The corresponding 95% CI for the *F*_1_ score difference was (0.037, 0.091), with an interval length of 0.054. In contrast, ignoring the correlation (using psF1 [14]) yielded a wider 95% CI of (0.032, 0.097), with a length of 0.065.

For future study planning, based on the estimated precision, sensitivity, and correlation values, a total of 741 skin cancer images, including 200 malignant cases, would be sufficient to achieve 80% power to detect an *F*_1_ score difference of 0.064 at a two-sided significance level of 0.05.

### 4.2 Breast cancer screening

This study evaluated whether an AI system could detect breast cancer from screening mammograms as accurately as, or better than, human radiologists. We applied psF1pair to compare the performance of the AI system and a human radiologist (the first reader) using the *F*_1_ score. The comparison was based on 25,522 screening mammograms from the UK 39-month dataset reported in Supplementary Table 1 of McKinney et al. [17], including 407 cancer cases and 25,115 non-cancer cases. The classification results are summarized in Table 2.

**Table 2.** Classification results for cancer and non-cancer cases in breast cancer screening study.

|  |  | **Non-cancer** |  | **Cancer** |  |
| --- | --- | --- | --- | --- | --- |
|  |  | **First reader** |  | **First reader** |  |
|  |  | Negative | Positive | Negative | Positive |
| **AI system** | Negative | 22,165 | 1,439 | 108 | 34 |
|  | Positive | 1,175 | 336 | 45 | 220 |
|  |  | **Consensus** |  | **Consensus** |  |
|  |  | Negative | Positive | Negative | Positive |
| **AI system** | Negative | 22,683 | 713 | 95 | 31 |
|  | Positive | 1,803 | 243 | 40 | 248 |

For the AI system and the first reader, the estimated precisions were 0.149 and 0.125 and the estimated sensitivity were 0.651 and 0.624, respectively. The resulting *F*_1_ scores were 0.243 and 0.209, yielding a statistically significant difference of 0.034 (95% CI: 0.018 to 0.051; p-value = 5.6 × 10^−5^). The results indicate that the AI system outperformed the first reader according to the *F*_1_ score metric. When the correlation between classifiers was ignored, the 95% CI widened to (0.011, 0.058).

The estimated correlations between the AI system and the first reader were 0.150 for cancer cases and 0.578 for non-cancer cases. Based on the estimated precision, sensitivity, and correlation values, a future study would require 12,761 screening mammograms, including 204 cancer cases, to achieve 80% power for detecting an *F*_1_ score difference of 0.034 at a two-sided significance level of 0.05.

Interestingly, when the AI system was compared with the consensus interpretation, using 25,856 screening mammograms (414 cancer cases and 25,442 non-cancer cases), the estimated precisions were 0.123 and 0.226, respectively, while the estimated sensitivity were 0.696 and 0.674. The resulting *F*_1_ scores were 0.210 for the AI system and 0.338 for the consensus interpretation. The *F*_1_ score difference was -0.129 (95% CI: -0.149 to -0.109; p-value < 1 × 10^−6^), indicating that the consensus interpretation performed better than the AI system.

## 5 Discussion

Reliable benchmarking of medical AI systems requires more than reporting point estimates of predictive performance. Confidence intervals, formal hypothesis testing, and prospective sample size determination are increasingly necessary for comparative validation studies. Because competing algorithms are typically evaluated on the same patient cohort, paired dependencies naturally arise and should be incorporated into statistical analyses. In this work, we developed psF1pair, a unified framework that addresses these challenges for *F*_1_ and *F_β_* performance metrics and enables inference and study design while accounting for paired classifier dependence.

Our simulation results demonstrate that the proposed confidence intervals for *F_β_* score difference achieve the desired nominal coverage, with accuracy improving as sample size increases. The empirical powers closely match the estimated powers, with differences within 3%, indicating that psF1pair provides reliable power calculations for study design. For hypothesis testing, when correlations are absent, psF1pair and the Wald-type Takahashi method perform best in scenarios with balanced precision and sensitivity or with equal sensitivity but distinct precision, while psF1pair performs better in the scenario with equal precision but distinct sensitivity. When correlations are present, psF1pair continues to outperform in the latter scenario. The Wald-type

Takahashi method achieves higher powers in the first two scenarios; however, it tends to inflate type I error rates when the number of positive cases is small to moderate, consistent with observations reported in Takahashi et al. [13].

psF1pair combines exact probability distributions for small sample sizes with asymptotic normal distributions for large sample sizes. Specifically, it applies the exact distribution approach for scenarios with a small number of positives and negatives and switches to the asymptotic approach for large sample sizes to reduce computational burden. By default, psF1pair automatically selects between these approaches based on whether the expected counts in all cells across both classifiers are at least five, similar to the rule of thumb used for chi-square approximations in contingency table analyses, thereby facilitating routine use in model evaluation and study-design applications. Our simulation study demonstrates that this automatic selection yields results nearly identical to those obtained using the exact distribution approach (Tables S13 and S14). In addition, psF1pair allows users to override the default behavior by specifying either the normal approximation (normal.approx = TRUE) or the exact distribution (normal.approx = FALSE) if desired.

Our simulation results indicate that higher correlations lead to greater statistical power and narrower interval lengths, thereby reducing required sample sizes. Therefore, specifying appropriate correlation values is an important consideration in study design. psF1pair can estimate correlations using pilot data, providing useful guidance for planning future large-scale studies. In the absence of pilot data, a conservative strategy is to assume zero correlations; although this results in a larger estimated sample size, it ensures that the desired power level is achieved. These findings highlight the importance of incorporating classifier dependence when designing comparative AI validation studies, as failure to do so may lead to unnecessarily large sample size requirements and inefficient study designs.

The proposed framework assumes independence across class instances and focuses on paired comparisons between two classifiers evaluated on the same dataset. Although these assumptions are common in validation studies, future work could investigate extensions to clustered or longitudinal data structures and comparisons involving multiple competing classifiers.

The increasing adoption of AI systems in radiology, pathology, dermatology, cancer screening, and clinical decision support underscores the importance of statistically rigorous evaluation methods. The results of this study demonstrate that psF1pair provides a practical framework for confidence interval estimation, hypothesis testing, and sample size determination while accounting for paired classifier dependence. By integrating these capabilities within a single framework, psF1pair may facilitate more efficient study planning, improve the rigor of comparative algorithm evaluations, and enhance the reproducibility of medical AI benchmarking studies.

## Data Availability

All data produced in the present work are contained in the manuscript.

https://github.com/cyhsuTN/psF1pair

## Data availability statement

The data used in this paper are included in Tables 1 and 2.

## Supplementary material

1. Online supplementary materials; 2. A freely available R package psF1pair at https://github.com/cyhsuTN/psF1pair

## References

1. Bhatt J, Jain S, Bhatia DD. Artificial intelligence in healthcare diagnosis: evidence-based recent advances and clinical implications. Sensors and Diagnostics. 2025;4:1047–1059.

2. Asif S, Wenhui Y, Tao Y, et al. An ensemble machine learning method for the prediction of heart disease. 2021 4th International Conference on Artificial Intelligence and Big Data (ICAIBD). 2021;98–103.

3. Ahmad GN, Fatima H, Ullah S, et al. Efficient medical diagnosis of human heart diseases using machine learning techniques with and without GridSearchCV. IEEE Access. 2022;10:80151–80173.

4. Bratchenko IA, Bratchenko LA, Khristoforova YA, et al. Classification of skin cancer using convolutional neural networks analysis of Raman Spectra. Comput Methods Programs Biomed. 2022;219:106755.

5. Shakeel PM, Burhanuddin M, Desa MI. Automatic lung cancer detection from CT image using improved deep neural network and ensemble classifier. Neural Comput Appl. 2022;34:9579–9592.

6. Bhattacharyya A, Bhaik D, Kumar S, et al. A deep learning based approach for automatic detection of COVID-19 cases using chest X-ray images. Biomed Signal Process Control. 2022;71:103182.

7. Cheng Y, Cheng R, Xu T, et al. Machine learning techniques applied to COVID-19 prediction: A systematic literature review. Bioengineering (Basel) 2025;12:514.

8. Ghanem M, Ghaith AK, El-Hajj VG, et al. Limitations in evaluating machine learning models for imbalanced binary outcome classification in spine surgery: a systematic review. Brain Sci. 2023;13:1723.

9. Norton ME, Jacobsson B, Swamy GK, et al. Cell-free DNA analysis for noninvasive examination of trisomy. N Engl J Med. 2015;372:1589–1597.

10. Asif S, Wenhui Y, ur-Rehman S, et al. Advancements and prospects of machine learning in medical diagnostics: unveiling the future of diagnostic precision. Archives of Computational Methods in Engineering. 2025;32:853–883.

11. Goutte C, Gaussier E. A probabilistic interpretation of precision, recall and f-score, with implication for evaluation. Advances in Information Retrieval. 2005;3408:345–359.

12. Lam KFY, Gopal V, Qian J. Confidence intervals for the F1 score: a comparison of four methods. arXiv. 2024. 10.48550/arXiv.2309.14621.

13. Takahashi K, Yamamoto K, Kuchiba A, et al. Hypothesis testing procedure for binary and multi-class F1-scores in the paired design. Stat Med. 2023;42:4177–4192.

14. Hsu CY, Liu Q, Shyr Y. A unified framework for statistical inference and power analysis of single and comparative Fβ scores. Stat Med. 2026; 45(10-12):e70557.

15. Flores P, Salicrú M, Sánchez-Pla A, et al. An equivalence test between features lists, based on the Sorensen–Dice index and the joint frequencies of GO term enrichment. BMC Bioinformatics. 2022;23:207.

16. Jinnai S, Yamazaki N, Hirano Y, et al. The development of a skin cancer classification system for pigmented skin lesions using deep learning. Biomolecules. 2020;10(8):1123.

17. McKinney, S.M., Sieniek, M., Godbole, V., Godwin, J., Antropova, N., Ashrafian, H. et al. International evaluation of an AI system for breast cancer screening. Nature. 2020;577:89–94.

