## Supplemental Materials for "A Unified Framework for Statistical Inference and Study Design of Comparative *F*_1_ and *F_β_* Scores under Paired Classifier Evaluations"

### Explicit form of the joint distribution of $\boldsymbol{Y}_i$

The joint probability mass function of $\boldsymbol{Y}_i = \boldsymbol{y}$ is given by

$$Ber_2(\boldsymbol{y}; p_1, p_2, \rho) = \sum_{\{\boldsymbol{x}:\, x_{10}+x_{11}=y_1, x_{01}+x_{11}=y_2\}} \pi_{00}^{x_{00}} \pi_{01}^{x_{01}} \pi_{10}^{x_{10}} \pi_{11}^{x_{11}},$$

where $\boldsymbol{x} = (x_{00}, x_{01}, x_{10}, x_{11})^T$ satisfies $x_{ll'} \geq 0$ for all $l, l' \in \{0,1\}$, and $x_{00} + x_{01} + x_{10} + x_{11} = 1$. $\boldsymbol{\pi} = (\pi_{00}, \pi_{01}, \pi_{10}, \pi_{11})^T$, with $\pi_{11} = \rho\sqrt{p_1(1-p_1)p_2(1-p_2)} + p_1 p_2$, $\pi_{10} = p_1 - \pi_{11}$, $\pi_{01} = p_2 - \pi_{11}$, and $\pi_{00} = 1 - \pi_{10} - \pi_{01} - \pi_{11}$.

### Explicit form of the joint distribution of the sum $\sum_i \boldsymbol{Y}_i$ (for $\boldsymbol{b}$ and $\boldsymbol{d}$)

The joint probability mass function of $\sum_{i=1}^{n} \boldsymbol{Y}_i = \boldsymbol{v}$ is given by

$$B_2(\boldsymbol{v}; n, p_1, p_2, \rho) = \sum_{\{\boldsymbol{z}:\, z_{10}+z_{11}=v_1, z_{01}+z_{11}=v_2\}} \frac{n!}{z_{00}!z_{01}!z_{10}!z_{11}!} \pi_{00}^{z_{00}} \pi_{01}^{z_{01}} \pi_{10}^{z_{10}} \pi_{11}^{z_{11}},$$

where $\boldsymbol{z} = (z_{00}, z_{01}, z_{10}, z_{11})^T$ satisfies $z_{ll'} \geq 0$ for all $l, l' \in \{0,1\}$, and $z_{00} + z_{01} + z_{10} + z_{11} = n$. $\boldsymbol{\pi} = (\pi_{00}, \pi_{01}, \pi_{10}, \pi_{11})^T$, with $\pi_{11} = \rho\sqrt{p_1(1-p_1)p_2(1-p_2)} + p_1 p_2$, $\pi_{10} = p_1 - \pi_{11}$, $\pi_{01} = p_2 - \pi_{11}$, and $\pi_{00} = 1 - \pi_{10} - \pi_{01} - \pi_{11}$.

### Bayesian estimation of Pearson correlation coefficients $\rho_N$ and $\rho_P$

The following outlines the procedure for estimating the Pearson correlation coefficients in a Bayesian framework:

1. Given observed data $\boldsymbol{Y}_i$, $i = 1, \ldots, N$, we define $z_{ll',Neg} = \sum_{i \in Neg} I(Y_{1i} = l,\ Y_{2i} = l')$ and $z_{ll',Pos} = \sum_{i \in Pos} I(Y_{1i} = l,\ Y_{2i} = l')$ for $l, l' \in \{0,1\}$. Let $\boldsymbol{z}_{Neg} = \left(z_{00,Neg}, z_{01,Neg}, z_{10,Neg}, z_{11,Neg}\right)^T$ and $\boldsymbol{z}_{Pos} = \left(z_{00,Pos}, z_{01,Pos}, z_{10,Pos}, z_{11,Pos}\right)^T$.
2. Assume that $\left(p_{00,c}, p_{01,c}, p_{10,c}, p_{11,c}\right)^T$ follow a Dirichlet prior with Jeffrey's non-informative parameters ($\boldsymbol{\alpha} = 0.5 \times \mathbf{1}$) for $c \in \{Neg, Pos\}$. Under this prior and the multinomial likelihoods of $\boldsymbol{z}_{Neg}$ and $\boldsymbol{z}_{Pos}$, the posteriors are Dirichlet with parameters $\boldsymbol{z}_{Neg} + 0.5 \times \mathbf{1}$ and $\boldsymbol{z}_{Pos} + 0.5 \times \mathbf{1}$, respectively.
3. Generate $\left(p_{00,c}, p_{01,c}, p_{10,c}, p_{11,c}\right)^T$ from the posterior Dirichlet distributions and calculate the Pearson correlation $r_c = (p_{11,c}p_{00,c} - p_{10,c}p_{01,c})/\sqrt{p_{1.,c}\, p_{0.,c}\, p_{.1,c}\, p_{.0,c}}$, where $p_{1.,c} = p_{11,c} + p_{10,c}$, $p_{.1,c} = p_{11,c} + p_{01,c}$, $p_{0.,c} = 1 - p_{1.,c}$, and $p_{.0,c} = 1 - p_{.1,c}$.
4. Repeat Step 3 a sufficiently large number of times and then obtain the posterior mean correlations, $\bar{r}_{Neg}$ and $\bar{r}_{Pos}$, which serve as Bayesian estimates of $\rho_N$ and $\rho_P$, respectively.

**Table S1**. Mean (standard deviation) of the lengths of 95% CIs for $F_{0.5}$ score difference across 1000 simulation runs. The second row presents coverage probability. Assume $\rho_N = \rho_P = \rho = 0$, 0.2, or 0.4.

| $\boldsymbol{p_{p1}}$ **= 0.6,** $\boldsymbol{p_{s1}}$ **= 0.8 (**$\boldsymbol{F_{0.5,1}}$ **= 0.632);** $\boldsymbol{p_{p2}}$ **= 0.7,** $\boldsymbol{p_{s2}}$ **= 0.9 (**$\boldsymbol{F_{0.5,2}}$ **= 0.733)** | | | |
|---|---|---|---|
| | $\boldsymbol{\rho = 0}$ | $\boldsymbol{\rho = 0.2}$ | $\boldsymbol{\rho = 0.4}$ |
| N=250, S=100 | .154 (.007)<br>.949 | .139 (.008)<br>.951 | .121 (.008)<br>.951 |
| N=100, S=40 | .244 (.016)<br>.949 | .220 (.017)<br>.954 | .193 (.018)<br>.950 |
| N=50, S=20 | .346 (.033)<br>.957 | .313 (.033)<br>.946 | .279 (.035)<br>.960 |
| N=1000, S=20 | .389 (.022)<br>.946 | .354 (.029)<br>.946 | .314 (.034)<br>.946 |
| $\boldsymbol{p_{p1} = p_{p2}}$ **= 0.6,** $\boldsymbol{p_{s1} = p_{s2}}$ **= 0.8 (**$\boldsymbol{F_{0.5,1} = F_{0.5,2}}$ **= 0.632)** | | | |
| | $\boldsymbol{\rho = 0}$ | $\boldsymbol{\rho = 0.2}$ | $\boldsymbol{\rho = 0.4}$ |
| N=250, S=100 | .156 (.008)<br>.954 | .139 (.008)<br>.954 | .122 (.008)<br>.956 |
| N=100, S=40 | .247 (.019)<br>.958 | .222 (.019)<br>.960 | .195 (.020)<br>.962 |
| N=50, S=20 | .354 (.035)<br>.956 | .323 (.039)<br>.968 | .283 (.038)<br>.966 |
| N=1000, S=20 | .403 (.024)<br>.954 | .366 (.031)<br>.951 | .321 (.035)<br>.938 |

**Table S2**. Mean (standard deviation) of the lengths of 95% CIs for $F_1$ score difference across 1000 simulation runs. The second row presents the coverage probability. Assume $\rho_N = \rho_P = \rho =$ 0, 0.2, or 0.4.

| $\boldsymbol{p_{p1}}$ = 0.6, $\boldsymbol{p_{s1}}$ = 0.8 ($\boldsymbol{F_{1,1}}$=0.686); $\boldsymbol{p_{p2}}$ = 0.7, $\boldsymbol{p_{s2}}$ = 0.9 ($\boldsymbol{F_{1,2}}$ = 0.788) | | | |
|---|---|---|---|
| | $\boldsymbol{\rho = 0}$ | $\boldsymbol{\rho = 0.2}$ | $\boldsymbol{\rho = 0.4}$ |
| N=250, S=100 | .146 (.008)<br>.953 | .132 (.008)<br>.948 | .116 (.008)<br>.953 |
| N=100, S=40 | .233 (.020)<br>.957 | .210 (.020)<br>.948 | .186 (.020)<br>.949 |
| N=50, S=20 | .330 (.040)<br>.957 | .300 (.039)<br>.954 | .269 (.039)<br>.950 |
| N=1000, S=20 | .356 (.035)<br>.942 | .325 (.037)<br>.953 | .291 (.036)<br>.952 |
| $\boldsymbol{p_{p1} = p_{p2}}$ = 0.6, $\boldsymbol{p_{s1} = p_{s2}}$ = 0.8 ($\boldsymbol{F_{1,1} = F_{1,2}}$ = 0.686) | | | |
| | $\boldsymbol{\rho = 0}$ | $\boldsymbol{\rho = 0.2}$ | $\boldsymbol{\rho = 0.4}$ |
| N=250, S=100 | .157 (.009)<br>.946 | .141 (.009)<br>.952 | .122 (.009)<br>.950 |
| N=100, S=40 | .249 (.022)<br>.957 | .224 (.022)<br>.956 | .197 (.022)<br>.954 |
| N=50, S=20 | .355 (.042)<br>.949 | .324 (.046)<br>.957 | .284 (.041)<br>.962 |
| N=1000, S=20 | .383 (.037)<br>.954 | .348 (.040)<br>.960 | .307 (.040)<br>.944 |

**Table S3**. Mean (standard deviation) of the lengths of 95% CIs for $F_2$ score difference across 1000 simulation runs. The second row presents coverage probability. Assume $\rho_N = \rho_P = \rho = 0$, 0.2, or 0.4.

| $p_{p1} = 0.6, p_{s1} = 0.8$ ($F_{2,1} = 0.750$); $p_{p2} = 0.7, p_{s2} = 0.9$ ($F_{2,2} = 0.851$) | | | |
|---|---|---|---|
| | $\rho = 0$ | $\rho = 0.2$ | $\rho = 0.4$ |
| N=250, S=100 | .162 (.013)<br>.946 | .145 (.013)<br>.949 | .128 (.013)<br>.950 |
| N=100, S=40 | .257 (.031)<br>.945 | .233 (.031)<br>.954 | .204 (.031)<br>.958 |
| N=50, S=20 | .361 (.061)<br>.940 | .329 (.061)<br>.949 | .297 (.058)<br>.939 |
| N=1000, S=20 | .370 (.061)<br>.950 | .339 (.061)<br>.946 | .306 (.058)<br>.942 |
| $p_{p1} = p_{p2} = 0.6, p_{s1} = p_{s2} = 0.8$ ($F_{2,1} = F_{2,2} = 0.750$) | | | |
| | $\rho = 0$ | $\rho = 0.2$ | $\rho = 0.4$ |
| N=250, S=100 | .182 (.013)<br>.952 | .163 (.014)<br>.945 | .141 (.014)<br>.954 |
| N=100, S=40 | .287 (.032)<br>.956 | .259 (.034)<br>.955 | .228 (.033)<br>.952 |
| N=50, S=20 | .405 (.062)<br>.939 | .369 (.067)<br>.951 | .326 (.062)<br>.962 |
| N=1000, S=20 | .413 (.062)<br>.956 | .376 (.064)<br>.961 | .333 (.062)<br>.960 |

**Table S4.** Power comparison across methods for the difference between two classifiers' $F_1$ scores at the significance level of 0.05. Assume $\rho_N = \rho_P = 0$.

| $\boldsymbol{p_{p1}}$ = 0.6, $\boldsymbol{p_{s1}}$ = 0.8 ($\boldsymbol{F_{1,1}}$ = 0.686); $\boldsymbol{p_{p2}}$ = 0.7, $\boldsymbol{p_{s2}}$ = 0.9 ($\boldsymbol{F_{1,2}}$ = 0.788) | | | | |
|---|---|---|---|---|
| | **psF1pair** | **Permutation** | **Takahashi (Wald)** | **Takahashi (Score)** |
| N=250, S=100 | .790 | .780 | .791 | .785 |
| N=100, S=40 | .398 | .370 | .389 | .380 |
| N=50, S=20 | .215 | .180 | .219 | .194 |
| N=1000, S=20 | .209 | .182 | .208 | .191 |
| $\boldsymbol{p_{p1}}$ = 0.6, $\boldsymbol{p_{s1}}$ = 0.8 ($\boldsymbol{F_{1,1}}$ = 0.686); $\boldsymbol{p_{p2}}$ = 0.8, $\boldsymbol{p_{s2}}$ = 0.8 ($\boldsymbol{F_{1,2}}$ = 0.800) | | | | |
| | **psF1pair** | **Permutation** | **Takahashi (Wald)** | **Takahashi (Score)** |
| N=250, S=100 | .797 | .788 | .802 | .790 |
| N=100, S=40 | .422 | .398 | .427 | .393 |
| N=50, S=20 | .227 | .220 | .231 | .200 |
| N=1000, S=20 | .223 | .212 | .228 | .198 |
| $\boldsymbol{p_{p1}}$ = 0.8, $\boldsymbol{p_{s1}}$ = 0.6 ($\boldsymbol{F_{1,1}}$=0.686); $\boldsymbol{p_{p2}}$ = 0.8, $\boldsymbol{p_{s2}}$ = 0.8 ($\boldsymbol{F_{1,2}}$ = 0.800) | | | | |
| | **psF1pair** | **Permutation** | **Takahashi (Wald)** | **Takahashi (Score)** |
| N=250, S=100 | .658 | .649 | .645 | .647 |
| N=100, S=40 | .314 | .285 | .288 | .283 |
| N=50, S=20 | .176 | .139 | .163 | .158 |
| N=1000, S=20 | .178 | .150 | .167 | .157 |

**Table S5.** Power comparison across methods for the difference between two classifiers' $F_1$ scores at the significance level of 0.05. Assume $\rho_N = \rho_P = 0.2$.

| $\boldsymbol{p_{p1}}$ = 0.6, $\boldsymbol{p_{s1}}$ = 0.8 ($\boldsymbol{F_{1,1}}$ = 0.686); $\boldsymbol{p_{p2}}$ = 0.7, $\boldsymbol{p_{s2}}$ = 0.9 ($\boldsymbol{F_{1,2}}$ = 0.788) | | | | |
|---|---|---|---|---|
| | **psF1pair** | **Permutation** | **Takahashi (Wald)** | **Takahashi (Score)** |
| N=250, S=100 | .870 | .860 | .870 | .868 |
| N=100, S=40 | .469 | .444 | .481 | .457 |
| N=50, S=20 | .265 | .229 | .268 | .249 |
| N=1000, S=20 | .239 | .213 | .251 | .225 |
| $\boldsymbol{p_{p1}}$=0.6, $\boldsymbol{p_{s1}}$=0.8 ($\boldsymbol{F_{1,1}}$ = 0.686); $\boldsymbol{p_{p2}}$=0.8, $\boldsymbol{p_{s2}}$ = 0.8 ($\boldsymbol{F_{1,2}}$ = 0.800) | | | | |
| | **psF1pair** | **Permutation** | **Takahashi (Wald)** | **Takahashi (Score)** |
| N=250, S=100 | .875 | .868 | .887 | .877 |
| N=100, S=40 | .509 | .516 | .549 | .506 |
| N=50, S=20 | .277 | .280 | .309 | .264 |
| N=1000, S=20 | .268 | .256 | .288 | .239 |
| $\boldsymbol{p_{p1}}$=0.8, $\boldsymbol{p_{s1}}$=0.6 ($\boldsymbol{F_{1,1}}$ = 0.686); $\boldsymbol{p_{p2}}$ = 0.8, $\boldsymbol{p_{s2}}$ = 0.8 ($\boldsymbol{F_{1,2}}$ = 0.800) | | | | |
| | **psF1pair** | **Permutation** | **Takahashi (Wald)** | **Takahashi (Score)** |
| N=250, S=100 | .743 | .733 | .723 | .726 |
| N=100, S=40 | .376 | .345 | .334 | .338 |
| N=50, S=20 | .192 | .159 | .181 | .179 |
| N=1000, S=20 | .208 | .171 | .190 | .190 |

**Table S6.** Power comparison across methods for the difference between two classifiers' $F_1$ scores at the significance level of 0.05. Assume $\rho_N$ = $\rho_P$ = 0.4.

| $p_{p1}$ = 0.6, $p_{s1}$ = 0.8 ($F_{1,1}$ = 0.686); $p_{p2}$ = 0.7, $p_{s2}$ = 0.9 ($F_{1,2}$ = 0.788) | | | | |
|---|---|---|---|---|
| | **psF1pair** | **Permutation** | **Takahashi (Wald)** | **Takahashi (Score)** |
| N=250, S=100 | .940 | .942 | .941 | .939 |
| N=100, S=40 | .591 | .579 | .608 | .585 |
| N=50, S=20 | .315 | .291 | .351 | .313 |
| N=1000, S=20 | .284 | .243 | .306 | .279 |
| **$p_{p1}$=0.6, $p_{s1}$=0.8 ($F_{1,1}$ = 0.686); $p_{p2}$=0.8, $p_{s2}$ = 0.8 ($F_{1,2}$ = 0.800)** | | | | |
| | **psF1pair** | **Permutation** | **Takahashi (Wald)** | **Takahashi (Score)** |
| N=250, S=100 | .944 | .940 | .959 | .954 |
| N=100, S=40 | .595 | .588 | .645 | .597 |
| N=50, S=20 | .347 | .353 | .414 | .343 |
| N=1000, S=20 | .341 | .326 | .390 | .324 |
| **$p_{p1}$=0.8, $p_{s1}$=0.6 ($F_{1,1}$ = 0.686); $p_{p2}$ = 0.8, $p_{s2}$ = 0.8 ($F_{1,2}$ = 0.800)** | | | | |
| | **psF1pair** | **Permutation** | **Takahashi (Wald)** | **Takahashi (Score)** |
| N=250, S=100 | .859 | .846 | .835 | .847 |
| N=100, S=40 | .480 | .431 | .415 | .443 |
| N=50, S=20 | .240 | .210 | .213 | .229 |
| N=1000, S=20 | .239 | .215 | .219 | .230 |

**Table S7**. Comparison of type I error rates across methods for the difference between two classifiers' $F_1$ scores at the significance level of 0.05. Assume $\rho_N = \rho_P = 0$.

| $\boldsymbol{p_{p1} = p_{p1} = 0.6, p_{s1} = p_{s2} = 0.8\ (F_{1,1} = F_{1,2} = 0.686)}$ | | | | |
|---|---|---|---|---|
| | **psF1pair** | **Permutation** | **Takahashi (Wald)** | **Takahashi (Score)** |
| N=250, S=100 | .055 | .049 | .053 | .053 |
| N=100, S=40 | .053 | .049 | .053 | .051 |
| N=50, S=20 | .053 | .050 | .055 | .053 |
| N=1000, S=20 | .052 | .044 | .049 | .048 |
| $\boldsymbol{p_{p1} = p_{p1} = 0.8, p_{s1} = p_{s2} = 0.6\ (F_{1,1} = F_{1,2} = 0.686)}$ | | | | |
| | **psF1pair** | **Permutation** | **Takahashi (Wald)** | **Takahashi (Score)** |
| N=250, S=100 | .049 | .044 | .050 | .046 |
| N=100, S=40 | .048 | .039 | .049 | .043 |
| N=50, S=20 | .050 | .042 | .056 | .047 |
| N=1000, S=20 | .054 | .044 | .055 | .049 |

**Table S8**. Comparison of type I error rates across methods for the difference between two classifiers' $F_1$ scores at the significance level of 0.05. Assume $\rho_N = \rho_P = 0.2$.

| $\boldsymbol{p_{p1} = p_{p1} = 0.6, p_{s1} = p_{s2} = 0.8\ (F_{1,1} = F_{1,2} = 0.686)}$ | | | | |
|---|---|---|---|---|
| | **psF1pair** | **Permutation** | **Takahashi (Wald)** | **Takahashi (Score)** |
| N=250, S=100 | .054 | .050 | .054 | .053 |
| N=100, S=40 | .050 | .039 | .053 | .049 |
| N=50, S=20 | .056 | .049 | .065 | .062 |
| N=1000, S=20 | .051 | .044 | .052 | .049 |
| $\boldsymbol{p_{p1} = p_{p1} = 0.8, p_{s1} = p_{s2} = 0.6\ (F_{1,1} = F_{1,2} = 0.686)}$ | | | | |
| | **psF1pair** | **Permutation** | **Takahashi (Wald)** | **Takahashi (Score)** |
| N=250, S=100 | .047 | .042 | .046 | .043 |
| N=100, S=40 | .053 | .045 | .053 | .050 |
| N=50, S=20 | .055 | .044 | .069 | .052 |
| N=1000, S=20 | .051 | .037 | .054 | .040 |

**Table S9**. Comparison of type I error rates across methods for the difference between two classifiers' $F_1$ scores at the significance level of 0.05. Assume $\rho_N = \rho_P = 0.4$.

| $\boldsymbol{p_{p1} = p_{p1} = 0.6, p_{s1} = p_{s2} = 0.8\ (F_{1,1} = F_{1,2} = 0.686)}$ | | | | |
|---|---|---|---|---|
| | **psF1pair** | **Permutation** | **Takahashi (Wald)** | **Takahashi (Score)** |
| N=250, S=100 | .053 | .055 | .054 | .051 |
| N=100, S=40 | .053 | .050 | .055 | .056 |
| N=50, S=20 | .047 | .039 | .060 | .059 |
| N=1000, S=20 | .049 | .047 | .054 | .059 |
| $\boldsymbol{p_{p1} = p_{p1} = 0.8, p_{s1} = p_{s2} = 0.6\ (F_{1,1} = F_{1,2} = 0.686)}$ | | | | |
| | **psF1pair** | **Permutation** | **Takahashi (Wald)** | **Takahashi (Score)** |
| N=250, S=100 | .049 | .047 | .052 | .047 |
| N=100, S=40 | .050 | .045 | .051 | .046 |
| N=50, S=20 | .053 | .045 | .066 | .065 |
| N=1000, S=20 | .051 | .041 | .062 | .060 |

**Table S10.** Empirical power (Estimated power calculated by psF1pair) for the $F_{0.5}$ score of a single classifier at the significance level of 0.05.

| **$p_{p1}$ = 0.6, $p_{s1}$ = 0.8 ($F_{0.5,1}$ = 0.632); $p_{p2}$ = 0.7, $p_{s2}$ = 0.9 ($F_{0.5,2}$ = 0.733)** | | | |
|---|---|---|---|
| | $\rho = 0$ | $\rho = 0.2$ | $\rho = 0.4$ |
| N=250, S=100 | .730 (.723) | .810 (.814) | .906 (.908) |
| N=100, S=40 | .360 (.358) | .420 (.431) | .544 (.542) |
| N=50, S=20 | .189 (.203) | .226 (.239) | .288 (.302) |
| N=1000, S=20 | .168 (.166) | .205 (.195) | .233 (.241) |
| **$p_{p1}$=0.6, $p_{s1}$=0.8 ($F_{0.5,1}$ = 0.632); $p_{p2}$=0.8, $p_{s2}$ = 0.8 ($F_{0.5,2}$ = 0.800)** | | | |
| | $\rho = 0$ | $\rho = 0.2$ | $\rho = 0.4$ |
| N=250, S=100 | .983 (.978) | .994 (.994) | 1.00 (.999) |
| N=100, S=40 | .712 (.696) | .815 (.793) | .905 (.896) |
| N=50, S=20 | .411 (.397) | .515 (.488) | .622 (.603) |
| N=1000, S=20 | .370 (.351) | .440 (.425) | .544 (.528) |
| **$p_{p1}$=0.8, $p_{s1}$=0.6 ($F_{0.5,1}$ = 0.750); $p_{p2}$ = 0.8, $p_{s2}$ = 0.8 ($F_{0.5,2}$ = 0.800)** | | | |
| | $\rho = 0$ | $\rho = 0.2$ | $\rho = 0.4$ |
| N=250, S=100 | .176 (.184) | .207 (.219) | .283 (.276) |
| N=100, S=40 | .113 (.100) | .115 (.114) | .138 (.138) |
| N=50, S=20 | .074 (.077) | .087 (.088) | .112 (.098) |
| N=1000, S=20 | .071 (.077) | .083 (.084) | .107 (.096) |

**Table S11.** Empirical power (Estimated power calculated by psF1pair) for the $F_1$ score of a single classifier at the significance level of 0.05.

| | | | |
|---|---|---|---|
| **$p_{p1}$ = 0.6, $p_{s1}$ = 0.8 ($F_{1,1}$ = 0.686); $p_{p2}$ = 0.7, $p_{s2}$ = 0.9 ($F_{1,2}$ = 0.788)** | | | |
| | $\rho = 0$ | $\rho = 0.2$ | $\rho = 0.4$ |
| N=250, S=100 | .790 (.771) | .870 (.855) | .940 (.935) |
| N=100, S=40 | .398 (.394) | .469 (.473) | .591 (.590) |
| N=50, S=20 | .215 (.219) | .265 (.263) | .315 (.329) |
| N=1000, S=20 | .209 (.195) | .239 (.232) | .284 (.291) |
| **$p_{p1}$=0.6, $p_{s1}$=0.8 ($F_{1,1}$ = 0.686); $p_{p2}$=0.8, $p_{s2}$ = 0.8 ($F_{1,2}$ = 0.800)** | | | |
| | $\rho = 0$ | $\rho = 0.2$ | $\rho = 0.4$ |
| N=250, S=100 | .797 (.788) | .875 (.871) | .944 (.947) |
| N=100, S=40 | .422 (.406) | .509 (.489) | .595 (.610) |
| N=50, S=20 | .227 (.220) | .277 (.267) | .347 (.342) |
| N=1000, S=20 | .223 (.208) | .268 (.250) | .341 (.318) |
| **$p_{p1}$=0.8, $p_{s1}$=0.6 ($F_{1,1}$ = 0.686); $p_{p2}$ = 0.8, $p_{s2}$ = 0.8 ($F_{1,2}$ = 0.800)** | | | |
| | $\rho = 0$ | $\rho = 0.2$ | $\rho = 0.4$ |
| N=250, S=100 | .658 (.644) | .743 (.737) | .859 (.846) |
| N=100, S=40 | .314 (.308) | .376 (.371) | .480 (.469) |
| N=50, S=20 | .176 (.177) | .192 (.208) | .240 (.260) |
| N=1000, S=20 | .178 (.174) | .208 (.205) | .239 (.254) |

**Table S12.** Empirical power (Estimated power calculated by psF1pair) for the $F_2$ score of a single classifier at the significance level of 0.05.

| $\boldsymbol{p_{p1}}$ = 0.6, $\boldsymbol{p_{s1}}$ = 0.8 ($\boldsymbol{F_{2,1}}$ = 0.750); $\boldsymbol{p_{p2}}$ = 0.7, $\boldsymbol{p_{s2}}$ = 0.9 ($\boldsymbol{F_{2,2}}$ = 0.851) | | | |
|---|---|---|---|
| | $\boldsymbol{\rho = 0}$ | $\boldsymbol{\rho = 0.2}$ | $\boldsymbol{\rho = 0.4}$ |
| N=250, S=100 | .682 (.679) | .782 (.773) | .886 (.877) |
| N=100, S=40 | .351 (.328) | .396 (.397) | .523 (.501) |
| N=50, S=20 | .179 (.183) | .222 (.217) | .257 (.271) |
| N=1000, S=20 | .198 (.180) | .221 (.212) | .275 (.264) |
| $\boldsymbol{p_{p1}}$=0.6, $\boldsymbol{p_{s1}}$=0.8 ($\boldsymbol{F_{2,1}}$ = 0.750); $\boldsymbol{p_{p2}}$=0.8, $\boldsymbol{p_{s2}}$ = 0.8 ($\boldsymbol{F_{2,2}}$ = 0.800) | | | |
| | $\boldsymbol{\rho = 0}$ | $\boldsymbol{\rho = 0.2}$ | $\boldsymbol{\rho = 0.4}$ |
| N=250, S=100 | .193 (.180) | .224 (.214) | .288 (.270) |
| N=100, S=40 | .108 (.098) | .115 (.111) | .127 (.133) |
| N=50, S=20 | .074 (.073) | .091 (.078) | .098 (.088) |
| N=1000, S=20 | .079 (.073) | .085 (.079) | .098 (.089) |
| $\boldsymbol{p_{p1}}$=0.8, $\boldsymbol{p_{s1}}$=0.6 ($\boldsymbol{F_{2,1}}$ = 0.632); $\boldsymbol{p_{p2}}$ = 0.8, $\boldsymbol{p_{s2}}$ = 0.8 ($\boldsymbol{F_{2,2}}$ = 0.800) | | | |
| | $\boldsymbol{\rho = 0}$ | $\boldsymbol{\rho = 0.2}$ | $\boldsymbol{\rho = 0.4}$ |
| N=250, S=100 | .830 (.834) | .919 (.905) | .974 (.965) |
| N=100, S=40 | .459 (.448) | .534 (.535) | .647 (.658) |
| N=50, S=20 | .271 (.251) | .300 (.296) | .363 (.384) |
| N=1000, S=20 | .255 (.251) | .297 (.297) | .379 (.384) |

**Table S13**. Comparison of the automatic approach, the normal approximation, and the exact approach in terms of mean (standard deviation) of the lengths of 95% CI and coverage probability (shown in the second rows), based on 1000 simulation runs for the F1 score difference. Assume $\rho_N = \rho_P = 0.2$.

| $\boldsymbol{p_{p1}}$ = 0.6, $\boldsymbol{p_{s1}}$ = 0.8 ($\boldsymbol{F_{1,1}}$ = 0.686); $\boldsymbol{p_{p2}}$ = 0.7, $\boldsymbol{p_{s2}}$ = 0.9 ($\boldsymbol{F_{1,2}}$ = 0.788) | | | |
|---|---|---|---|
| | **auto** | **TRUE** | **FALSE** |
| N=250, S=100 | .132 (.008)<br>.948 | .132 (.008)<br>.948 | .132 (.008)<br>.948 |
| N=100, S=40 | .210 (.020)<br>.948 | .210 (.020)<br>.948 | .211 (.020)<br>.949 |
| N=50, S=20 | .300 (.039)<br>.954 | .296 (.038)<br>.958 | .300 (.039)<br>.954 |
| N=1000, S=20 | .325 (.037)<br>.953 | .323 (.036)<br>.955 | .325 (.037)<br>.953 |
| $\boldsymbol{p_{p1} = p_{p2}}$ = 0.6, $\boldsymbol{p_{s1} = p_{s2}}$ = 0.8 ($\boldsymbol{F_{1,1} = F_{1,2}}$ = 0.686) | | | |
| | **auto** | **TRUE** | **FALSE** |
| N=250, S=100 | .141 (.009)<br>.952 | .141 (.009)<br>.952 | .141 (.009)<br>.952 |
| N=100, S=40 | .224 (.022)<br>.956 | .224 (.023)<br>.956 | .226 (.023)<br>.954 |
| N=50, S=20 | .324 (.046)<br>.957 | .320 (.046)<br>.959 | .325 (.047)<br>.957 |
| N=1000, S=20 | .348 (.040)<br>.960 | .346 (.039)<br>.957 | .349 (.040)<br>.960 |

**Table S14**. Comparison of the automatic approach, the normal approximation, and the exact approach in terms of power and type I error rate, based on 1000 simulation runs. Assume $\rho_N = \rho_P = 0.2$.

| $p_{p1} = 0.6, p_{s1} = 0.8$ $(F_{1,1} = 0.686)$; $p_{p2} = 0.7, p_{s2} = 0.9$ $(F_{1,2} = 0.788)$ | | | |
|---|---|---|---|
| | **auto** | **TRUE** | **FALSE** |
| N=250, S=100 | .870 | .870 | .870 |
| N=100, S=40 | .469 | .473 | .467 |
| N=50, S=20 | .265 | .276 | .263 |
| N=1000, S=20 | .239 | .240 | .239 |
| $p_{p1} = p_{p2} = 0.6, p_{s1} = p_{s2} = 0.8$ $(F_{1,1} = F_{1,2} = 0.686)$ | | | |
| | **auto** | **TRUE** | **FALSE** |
| N=250, S=100 | .054 | .054 | .053 |
| N=100, S=40 | .050 | .050 | .050 |
| N=50, S=20 | .056 | .059 | .053 |
| N=1000, S=20 | .050 | .051 | .050 |
